# Rare variants found in multiplex families with orofacial clefts: Does expanding the phenotype make a difference?

**DOI:** 10.1101/2023.02.01.23285340

**Authors:** Kimberly K. Diaz Perez, Sydney Chung, S. Taylor Head, Michael P. Epstein, Jacqueline T. Hecht, George L. Wehby, Seth M. Weinberg, Jeffrey C. Murray, Mary L. Marazita, Elizabeth J. Leslie

## Abstract

Whole-exome sequencing (WES) is now a relatively straightforward process to identify causal variants in Mendelian disorders. However, the same is not true for WES in families where the inheritance patterns are less clear, and a complex etiology is suspected. Orofacial clefts (OFCs) are highly heritable birth defects with both Mendelian and complex etiologies. The phenotypic spectrum of OFCs may include overt clefts and several subclinical phenotypes, such as discontinuities in the *orbicularis oris* muscle (OOM) in the upper lip, velopharyngeal insufficiency (VPI), microform clefts or bifid uvulas. We hypothesize that expanding the OFC phenotype to include these phenotypes can clarify inheritance patterns in multiplex families, making them appear more Mendelian. We performed whole-exome sequencing to find rare, likely causal genetic variants in 31 multiplex OFC families, which included families with multiple individuals with OFCs and individuals with subclinical phenotypes. We identified likely causal variants in *COL11A2, IRF6, KLF4, SHROOM3, SMC3, TP63*, and *TBX3* in seven families. Although we did not find clear evidence supporting the subclinical phenotype hypothesis, our findings support a role for rare variants in the etiology of OFCs.

## INTRODUCTION

Orofacial clefts (OFCs) represent a human disorder where rare and common variant studies have been successful (Leslie, 2022). OFCs are common birth defects (affecting 1/1000 live births worldwide) that occur on an etiological spectrum that includes Mendelian genetic causes as well as environmental causes, such as exposure to teratogens during pregnancy (Garland et al., 2020). However, most OFCs are thought to occur as complex disorders resulting from the interaction of multiple genetic risk factors and environmental influences (Beaty et al., 2016). Mendelian forms of OFCs are often syndromes that can include non-cleft phenotypes in some affecteds as opposed to isolated, non-syndromic cases with accompanying additional clinical features (Dixon et al., 2011). It is now clear from multiple studies that non-syndromic and syndromic forms of OFCs have overlapping etiological spectrums (Basha et al., 2018; Leslie, 2022). One hypothesis arising from sequencing studies suggests that pathogenic variants causing syndromic OFCs tend to be deleterious exonic variants in genes involved in craniofacial development (Kondo et al., 2002; Peyrard-Janvid et al., 2014) whereas variants associated with non-syndromic OFCs may have less severe effects on protein function or occur in regulatory variants of the same genes (Leslie et al., 2016; Rahimov et al., 2008; Zucchero et al., 2004). However, the genetic mechanisms for risk in non-syndromic OFCs are varied and include complex/oligogenic/multigenic mechanisms (Alade et al., 2022; Stanier & Moore, 2004), Mendelian variants (Cox et al., 2018; Liu et al., 2017), and de *novo* mutations (Awotoye et al., 2022; Bishop et al., 2020); but much of the risk for OFCs is still unknown.

Genetic studies of non-syndromic OFCs have recently favored genome-wide association studies (GWAS) and over 15 GWAS or meta-analyses have cumulatively identified over 50 associated genes or loci (Alade et al., 2022; Birnbaum et al., 2009; Leslie, 2022; Leslie et al., 2016; Mangold et al., 2010; Yu et al., 2017). These loci are estimated to account for only ∼20-25% of the known heritable risk of OFCs, leaving a substantial portion of risk variants unaccounted for (Alade et al., 2022; Leslie, 2022). Decreases in the cost of sequencing that allow for far larger sample sizes to be studied have facilitated a shift toward the analysis of rare genetic variation as a possible source of OFC risk, as they are hypothesized to have larger effect sizes compared to common variants (Kryukov et al., 2007).

One approach to identify rare variants is to focus on family-based study designs as rare variants with large effects might segregate with OFCs in multiplex families. In support of this, Bureau et al. (2014) and Cox et al. (2018) identified rare, “likely pathogenic” variants shared by affected relatives that segregated in a dominant manner within ostensibly non-syndromic OFC families. Basha et al. (2018) estimated that rare “likely pathogenic” variants in genes associated with OFC syndromes could be identified in ∼10% of multiplex non-syndromic OFC families.

Approximately 15% of families with non-syndromic OFCs are multiplex, but the pattern of affected relatives does not always follow classic Mendelian patterns. Imposing a Mendelian structure on these families would require high levels of incomplete penetrance as there can be multiple unaffected individuals linking the affected individuals (Kingdom & Wright, 2022). We have previously hypothesized that this “incomplete penetrance” could be explained by the phenotypic misclassification of individuals who lack overt OFCs but have subclinical phenotypes (Marazita, 2012; Weinberg et al., 2006). Under this hypothesis, individuals manifesting these subclinical cleft features could represent “genetic carriers” who, because the phenotype is so subtle, are mischaracterized as unaffected. This expanded phenotypic spectrum of OFCs includes subclinical phenotypes such as discontinuities in the *orbicularis oris muscle* (OOM), velopharyngeal insufficiency (VPI), or mild phenotypes, such as bifid uvula (Weinberg et al., 2006). OOM discontinuities are subepithelial defects of the muscle surrounding the upper lip and are only detected through ultrasonography. Similarly, VPI is not readily observable and occurs when the muscular valve between the oral and nasal cavity fails to close, resulting in hypernasal speech and phonation challenges (Weinberg et al., 2006). These subclinical phenotypes are hypothesized to be mild forms of OFCs in part because they have been observed at higher frequencies in apparently unaffected individuals from OFC families compared to controls (Neiswanger et al., 2007; Weinberg et al., 2006). Here, we hypothesize that including these subclinical phenotypes could clarify the inheritance patterns in multiplex OFC families and help the identification of genetic risk factors segregating in these families.

Therefore, we aimed to investigate rare coding variants in multiplex OFC families with whole-exome sequencing by testing two complementary hypotheses. First, we hypothesized that multiplex families with inheritance patterns consistent with a Mendelian mechanism would segregate private, rare variants among affected individuals. Second, we hypothesized that subclinical OFC phenotypes would increase support for specific inheritance patterns and that likely causal variants would segregate among individuals with either overt phenotypes or subclinical phenotypes.

## METHODS

### Cohort Information

This study cohort consists of 31 families from national and international recruitment sites in the United States (Colorado, Iowa, Pennsylvania, Texas) (N=13), Europe (Hungary) (N=2), Asia (China, India, Philippines) (N=13), and Central America (Guatemala) (N=3) originally recruited for the Pittsburgh Orofacial Cleft Study at the University of Pittsburgh. All participants provided informed consent; the study was approved by the IRB at the University of Pittsburgh and local recruiting sites. We selected apparently non-syndromic OFC families for sequencing if they met the criteria for one of the following groups: (I) OFC multiplex families: characterized by the presence of at least one set of second degree or closer relative pairs where each member had an OFC (CL, CLP, or CP) and lack sequenced individuals with subclinical phenotypes (N=12); (II) multiplex families with subclinical phenotypes: contains multiple sequenced affected individuals as well as relatives with at least one subclinical phenotype (N=19). Most families had demographic and medical histories as well as photographs of the study participants. A total of 150 individuals (75 males, 75 females) with sufficient DNA quantities were sequenced (Supplemental Table 1).

### Sequencing

Whole-exome sequencing was performed using the Agilent SureSelectXT HumanAllExon V6 + UTR S07604624 exome capture kit at the Center for Inherited Disease Research. A low-input library prep protocol developed at CIDR was performed (Marosy et al., 2017). Libraries were prepared from 50ng of genomic DNA, sheared for 80s using the Covaris E220 instrument (Covaris). The KAPA Hyper prep kit was used to process the sheared DNA into amplified dual indexed adapter-ligated fragments. 750ng of the amplified library was used in an enrichment reaction following Agilent protocols. Libraries were sequenced on the NovaSeq 6000 platform with onboard clustering using 125 base pairs paired-end runs and sequencing chemistry kit NovaSeq 6000 S4 Reagent Kit v1.

### Variant Calling and Quality Control

Fastq files were aligned with BWA-MEM version 0.7.15 to the 1000 genomes phase 2 (GRCh37) human genome reference (Li, 2013). Duplicate molecules were flagged with Picard version 2.17.0. Base call quality score recalibration and binning (2,10,20,30) were performed using the Genome Analysis Toolkit (GATK) version v4.0.1.1 (McKenna et al., 2010). Cram files were generated using SAMTools version 1.5. GATK’s reference confidence model workflow was used to perform joint sample genotyping using GATK version 3.7. Briefly, this workflow entails: 1) Producing a gVCF (genomic Variant Call Format (VCF)) for each sample individually using Haplotype Caller (--emitRefConfidence GVCF) and –max_alternate_alleles was set to 3 to all bait intervals to generate likelihoods that the sites are homozygote reference or not, and 2) Joint genotyping the single sample gVCFs together with GenotypeGVCFs to produce a multi-sample VCF file. Variant filtering was done using the Variant Quality Score Recalibration (VQSR) method (DePristo et al., 2011). For single-nucleotide variants (SNVs), the annotations of MQRankSum, QD, FS, ReadPosRankSum, MQ, and SOR were used in the adaptive error model. HapMap3.3, Omni2.5, and 1000G phase high confidence SNP calls were used as training sets with HapMap3.3 and Omni2.5 used as the truth set. SNVs were filtered to obtain all variants up to the 99.5^th^ percentile of truth sites (0.5% false negative rate). For indels, the annotations of FS, ReadPosRankSum, MQRankSum, QD, and SOR were used in the adaptive error model (4 maximum Gaussians allowed). A set of curated indels obtained from the GATK resource bundle (Mills_and_1000G_gold_standard.indels.b37.vcf) were used as training and truth sites. Indels were filtered to obtain all variants up to the 99^th^ percentile of truth sites (1% false negative rate). Prior to the analysis, additional filters on genotype calls were applied based on a read depth ≥ 15 and genotype quality ≥ 20 via VCFtools (version 0.1.13).

### Variant Filtering

All variants within each family were annotated using Bystro Genomics (Kotlar et al., 2018), an in-house variant annotation and filtering tool, and VarSeq v2.2.5 (Golden Helix, Inc., Bozeman, MT). We retained and analyzed variants that met the following criteria: 1) exonic, 2) missense, nonsense, frameshift, and canonical splice variants, and 3) a global minor allele frequency (MAF) ≤ 0.5% in the Genome Aggregation Database (gnomAD) exomes and genomes v.2 (Karczewski et al., 2020). We also considered predictors of missense pathogenicity using various *in silico* tools, such as CADD scores (Rentzsch et al., 2018) and gene tolerance to variation metrics from gnomAD (Karczewski et al., 2020). Gene tolerance measures included Z-scores for missense variants, the probability of being loss-of-function intolerant (pLI) (Lek et al., 2016), and loss-of-function observed/expected upper bound fraction (LOEUF) for loss-of-function variants from gnomAD (Karczewski et al., 2020).

### Single-Family Segregation Analyses

For single-family analyses, we defined individuals with either an overt cleft or a subclinical phenotype as “affected”. In families with an apparent dominant inheritance pattern, we analyzed heterozygous variants shared among all affected individuals. In families with an apparent recessive mode of inheritance, we analyzed compound heterozygous variants or homozygous variants in the affected individuals and both parents had to be carriers for the variant. For multiplex families with only overt clefts, we allowed variants to be present in their unaffected relatives to allow for an incompletely penetrant model. After filtering for variants using the criteria noted above, we performed literature searches using databases, such as ClinVar (Landrum et al., 2018) and Online Mendelian Inheritance in Man (OMIM) (Hamosh et al., 2005), to support the plausibility of the variant and the gene to cause OFCs or a craniofacial phenotype.

### Mixed Model Linear Regression

We conducted linear mixed-effect models to compare the number of variants in individuals with OFCs and subclinical phenotypes. We utilized the “lme4” (version 1.1-29) package (Bates et al., 2015) along with the “afex” package (version 1.2-0) (Henrik et al., 2022) in R (version 3.6.3) (Team, 2021). We computed the number of heterozygous rare, protein-altering variants (MAF ≤ 0.5%) for each sample in protein-coding genes and OFC genes. We considered affection status (presence of an OFC) and the presence of subclinical phenotypes as indicator variables. We added a family-specific random intercept to account for relatedness within families. For models considering variants in OFC genes, we utilized a gene list comprised of 418 genes previously associated with craniofacial development and OFCs. These were compiled from four sources, including OMIM (Amberger et al., 2015), the PreventionGenetics Cleft Lip/Cleft Palate Panel (PreventionGenetics, Marshfield, WI), the Genomics England PanelApp Clefting Panel (v2.2, March 2020; (Martin et al., 2019)), and an additional set of literature-curated genes (more information on this gene list can be found in Diaz Perez et al. (2022). We considered a significance level for both the “all genes” and “OFC genes” at P < 0.05.

## RESULTS

We filtered for rare variants in protein-coding regions (MAF ≤ 0.5%) in 31 total families and identified an average of 195 variants in affected individuals (range 12 to 655 variants) per family.

### Variants in Families with Overt Clefts

In the 12 families that only had individuals with overt clefts, we identified likely causal variants in three families (3/12, 25%).

Family 1: Family 1 was a Guatemalan family comprised of four siblings with CLP, four unaffected siblings, and unaffected parents. All four siblings shared a novel 1 bp deletion in *TP63* leading to a frameshift (NM_003722.5: c.1606delC; NP_003713.3: p.His536Thrfs*18). The variant was inherited from the unaffected mother and was also found in their unaffected sister (Figure 1A). *TP63* is highly intolerant to loss-of-function variation (pLI = 1, LOEUF = 0.27) and the variant was not present in gnomAD. Heterozygous missense mutations in *TP63* cause allelic syndromes impacting the face and/or limbs including ectrodactyly, ectodermal dysplasia and CL/P (EEC) syndrome, and ankyloblepharon-ectodermal defects-CL/P (AEC) syndrome. Deletions and frameshift variants in *TP63* have recently been identified in non-syndromic OFC families (Khandelwal et al., 2019). Interestingly, like Family 1, most of the published variants were inherited from unaffected parents, suggesting an incompletely penetrant effect for truncating mutations in *TP63*. To support the initial diagnosis of non-syndromic OFC, we examined photographs of the family which did not reveal any evidence of ectodermal dysplasia or limb defects associated with *TP63*-associated syndromes. Because all affected individuals were male and there was no male-to-male transmission that would rule out an X-linked inheritance model, we also examined genes on the X chromosome. There was one variant in *SEPTIN6* (NP_665798.1:p.Ser408Cys) that was heterozygous in the mother and was transmitted to the affected male offspring but not the unaffected male or unaffected sister. However, it is found as a hemizygous variant in 23 males in the Latino/Admixed American population in gnomAD (0.3% allele frequency) and this makes it a less compelling candidate than the novel, truncation variant in *TP63*.

**Figure 1.**
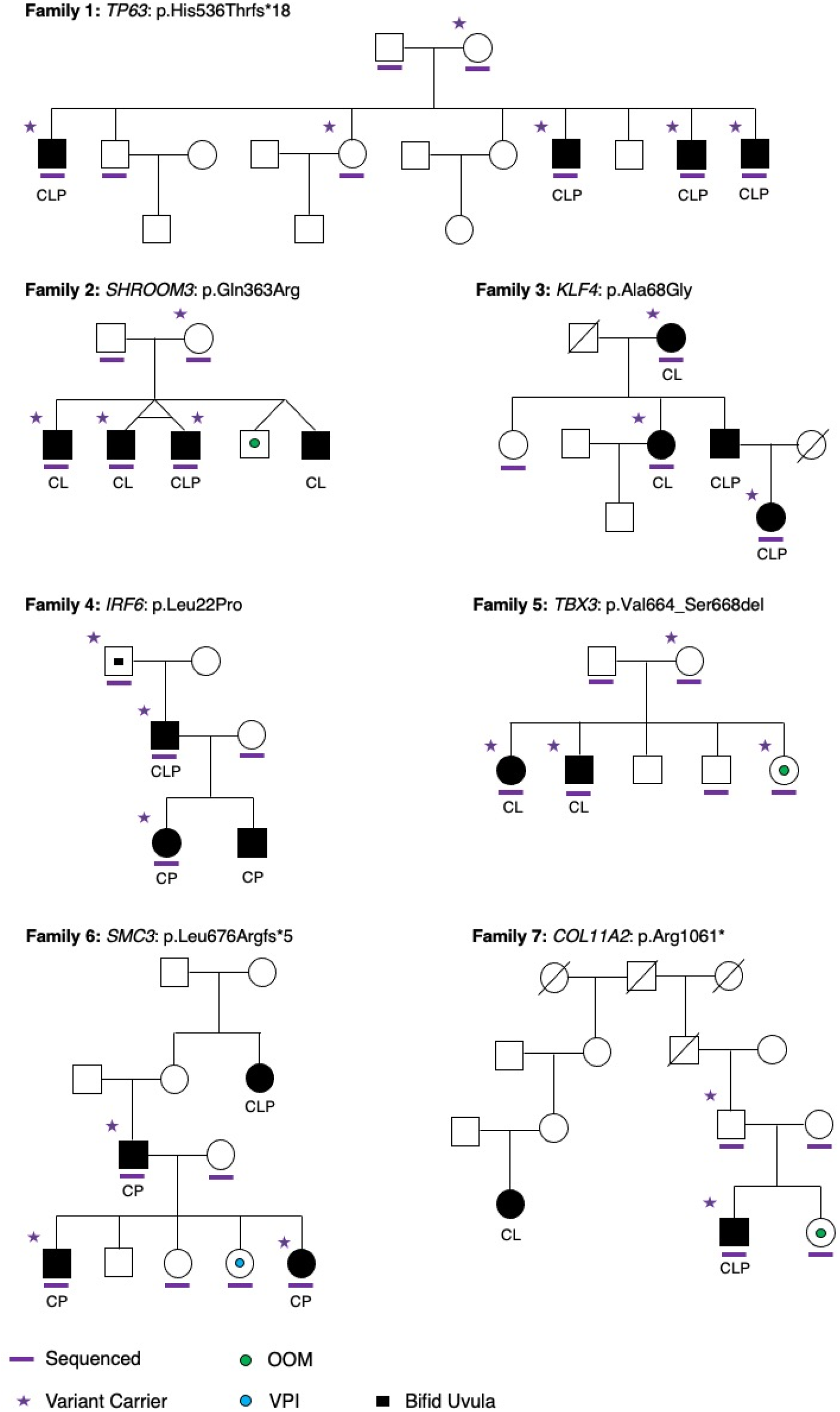
Likely Causal Variants in Multiplex OFC Families. We found seven likely causal variants in (A) *TP63*, (B) *SHROOM3*, (C) *KLF4*, (D) *IRF6*, (E) *TBX3*, (F), *SMC3*, and (G) C*OL11A2*. Sex symbols with solid black indicate the phenotype of the individual: CL (cleft lip), CP (cleft palate), and CLP (cleft lip and palate). The symbol with a green circle represents the individuals with discontinuities in the orbicularis oris muscle (OOM), the blue circle represents individuals with velopharyngeal insufficiency (VPI), and the black solid square inside the symbol indicates the sample had a bifid uvula. The purple solid lines indicate individuals with whole-exome data while purple solid stars indicate variant carriers.

Family 2: We identified a heterozygous missense mutation in *SHROOM3* (NM_020859.4: c.1088A>G; NP_065910.3: p.Gln363Arg) in a family from the United States that was shared between a set of monozygotic twins, one with CL and the other with CLP, and their brother with CL, and was transmitted from their unaffected mother (Figure 1B). *SHROOM3* is associated with the cytoskeleton, and it is important for neural tube morphogenesis (Das et al., 2014; Hildebrand & Soriano, 1999). *SHROOM3* has been previously associated with OFCs through genome-wide association studies and rare, de novo mutations in OFC trios (Bishop et al., 2020; Copp & Greene, 2013; Leslie et al., 2017; Ray et al., 2021). Moreover, mouse mutants of Shroom3 have been shown to exhibit highly penetrant craniofacial malformations, including exencephaly and facial clefting (Hildebrand & Soriano, 1999).

Family 3: We found a missense substitution in *KLF4* (NM_004235.6: c.203C>G; NP_004226.3: p.Ala68Gly) in a three-generation Chinese family that segregated among all three affected individuals with CL or CLP and was absent from the sequenced unaffected individual (Figure 1C). *KLF4* is a transcription factor involved in the differentiation of the epidermis (Segre et al., 1999). The expression of *KLF4* is directly regulated by IRF6, a key OFC-associated gene, in the oral epithelium during periderm differentiation in zebrafish (Liu et al., 2016). Functional zebrafish studies of rare missense mutations in *KLF4* in non-syndromic OFC cases found alterations in the differentiation of the periderm, indicating that rare variants in *KLF4* may increase the risk for OFCs (Liu et al., 2020).

### Variants in Families with Overt Clefts and Subclinical Phenotypes

We evaluated 19 multiplex OFC families with at least one sequenced individual with a subclinical phenotype. We found likely causal variants in four families (4/19, 21%) (Figure 2).

**Figure 2.**
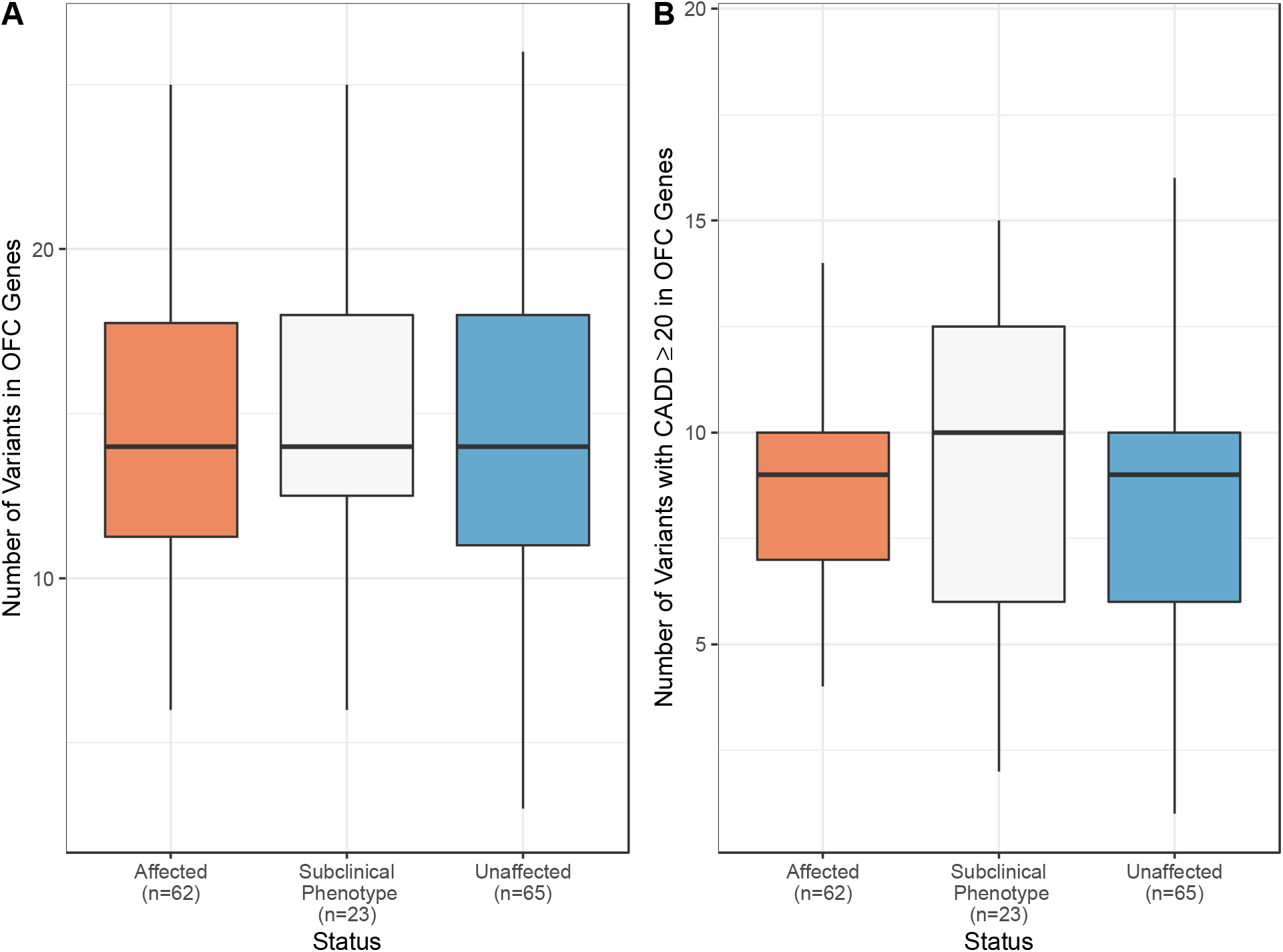
Number of Variants in OFC Genes per Family Within Groups. The number of variants in genes associated with OFCs per person (A) overall and (B) variants with a CADD ≥ 20 across affection status, including affected individuals (n=62, orange), individuals with subclinical phenotypes (n=23, gray) and unaffected (n=65, blue) individuals.

Family 4: We identified a novel missense mutation in *IRF6* (NM_006147.4: c.65T>C; NP_006138.1: p.Leu22Pro) in a three-generation pedigree from Hungary (Figure 1E). This substitution is located in the DNA-binding domain of the IRF6 protein and has been previously reported in Van der Woude syndrome (VWS) (Ghassibé et al., 2004). The variant was transmitted from the paternal grandfather, who had a bifid uvula, missing teeth, and syndactyly of the hands and feet. The proband’s father, who had CLP and missing teeth, also had the missense variant. Lip pits, one of the diagnostic criteria for VWS, were not reported. Ink lip prints (Neiswanger et al., 2009), but not photographs, were collected; however, it is not possible to conclusively confirm the presence or absence of lip pits from these prints.

Family 5: In Family 5 from the Philippines, we found a rare in-frame deletion (NM_005996.4: c.1991_2005delTGGCAGTGGACTCGG; NP_005987.3: p.Val664_Ser668del) in *TBX3* (Figure 1F). The deletion was transmitted from the unaffected mother and was present in two affected individuals and a sibling with the OOM phenotype, but not their unaffected siblings. Heterozygous truncation mutations in *TBX3* mutations cause Ulnar-mammary syndrome, characterized by mammary gland hypoplasia and upper limb defects. The proband is a short (5’ 2”), but not obese (<100 lbs), female with a missing lateral right incisor and unilateral CL. At the time of enrollment, no developmental delays or other structural anomalies were reported. The mother reported a history of miscarriage but did not any major medical conditions or structural anomalies; a limited craniofacial physical exam by research staff reported buccal frenula and a high arched palate. Although OFC rarely occurs in Ulnar-mammary syndrome, inactivation of *TBX3* in the neural crest in mice leads to postnatal death and a highly penetrant cleft palate (López et al., 2018).

Family 6: We identified a novel 32-base pair deletion in *SMC3* (NM_005445.4: c.2019_2050del; NP_005436.1: p.Leu676Argfs*5) in this family from the United States (Figure 1G). The variant was shared between the three affected individuals with CP but was not present in the sibling with VPI (Figure 1E). The deletion was paternally inherited and the father’s unsequenced aunt had CLP. Mutations in *SMC3* cause Cornelia de Lange (CdL) syndrome; however, this family did not have any additional structural anomalies, intellectual disability, or craniofacial features (e.g., microcephaly, arched eyebrows) that are characteristic of CdL (Gil-Rodriguez et al., 2015; Kline et al., 2007). This family illustrates that the inclusion of subclinical phenotypes could lead to false negatives should the causal variant for OFCs not be also causal for the subclinical phenotype.

Family 7: We identified a nonsense mutation in COL11A2 (NM_080680.3: c.3181C>T; NP_542411.2: p.Arg1061*) in a family from the United States that was transmitted to the proband with CLP from his unaffected father but was not present in his sibling with an OOM defect (Figure 1H). *COL11A2* is associated with autosomal dominant and recessive forms of Fibrochondrogenesis and Otospondylomegaepiphyseal Dysplasia (also known as non-ocular Stickler syndrome), the latter of which sometimes includes cleft palate (van Steensel et al., 1997; Vikkula et al., 1995). *COL11A2* has also been associated with non-syndromic OFCs through common variants (Nikopensius et al., 2010).

### Quantitative Variant Analysis

In most families, we were not able to identify a single causal variant, but we did observe many compelling missense variants in genes associated with craniofacial development (Supplemental Table 2). We hypothesized that individuals with overt clefts might have a higher number of such variants compared to their relatives with subclinical phenotypes. Using a curated list of 418 genes, we first calculated the number of rare (MAF ≤ 0.5%), protein-altering variants in individuals with overt clefts or subclinical phenotypes. We found fewer variants in individuals with OFCs (an average of 14.3 variants per person) than in individuals with subclinical phenotypes (an average of 15 variants per person) (Figure 2A). After adjusting for affection status and relatedness, there was no difference in the number of variants in all protein-coding genes (p=0.46) or OFC genes (p=0.64). The same was true when restricting to rare variants with a CADD score ≥ 20 (Figure 2B; p=0.27 for protein-coding genes and p=0.44 for OFC genes).

## DISCUSSION

In this study, we aimed to investigate the contribution of rare variants in the genetic etiology of OFCs by sequencing 31 multiplex families with overt OFCs with or without subclinical phenotypes. Our “hit” rate was ∼21-25% for both families with individuals with subclinical phenotypes and families with overt OFCs only, which is higher than the 10% reported by Basha et al. (2018), but is not statistically different (p=0.21, Fisher’s exact test). Our higher rate may be explained by the smaller sample size but there were also differences in the selection of families and the analysis pipeline. One of our families had an IRF6 mutation, but this family (and others like them) would have been excluded from the Basha et al. study, which were drawn from a database prescreened for IRF6 mutations. Basha et al. also focused their analysis on a subset of 500 genes plausibly involved in OFCs.

Rare variants in *BMP4* were previously reported to be associated with overt clefts and OOM defects; however, *BMP4* variants were not found among the candidate variants in this study (Suzuki et al., 2009). In fact, we did not detect strong evidence to suggest that the inclusion of subclinical phenotypes facilitates gene discovery. Given the small sample sizes in this study, our evidence supporting a common etiology for subclinical phenotypes and overt OFCs is only anecdotal. Additional genetic studies need to be conducted in larger and more phenotypically homogeneous samples to determine the utility of subclinical phenotypes for gene discovery.

Four variants were transmitted from unaffected parents. One explanation for incomplete penetrance of a variant is mosaicism in the transmitting parent (Kingdom & Wright, 2022). We have limited ability to detect mosaicism with a single tissue source and standard exome sequencing, but nonetheless did not find evidence of mosaicism in the parental samples based on the allele balance (43.2-52.4% alternate alleles). It is also possible the effect of the variant is modified by as-yet unknown environmental exposures or additional genetic risk factors, which could influence the expression of OFCs (Beames & Lipinski, 2020; Carlson et al., 2017). Similar explanations (e.g. mosaicism, modifiers, or stochastic events) may explain the variable expressivity of overt and more mild forms of OFCs observed in these families. More work is needed to test the hypothesis that OFCs and subclinical phenotypes share an etiology and to determine the impact of rare genetic variation in the etiology of OFCs.

Overall, our results provide further evidence of the Mendelian transmission of rare coding variants in non-syndromic multiplex OFC families. Similar to the findings of Basha et al. (2018), Bishop et al. (2020), and others, this work provides evidence that individuals and families with apparently non-syndromic OFCs may have rare coding variants in genes associated with syndromic OFCs. These results can provide support for the recommendation to offer diagnostic genetic testing to families with apparently non-syndromic OFCs and a positive family history. We note, however, that the number of affected family members and the family structure should be carefully considered. Many of our families were relatively small and not all affected or informative individuals had DNA available or were successfully sequenced, limiting our ability to narrow the list of candidate variants. In this study, we found most likely causal variants in families with at least three affected individuals. Specific recommendations for diagnostic testing will continue to evolve as more data on the contribution of rare variants to both isolated and familial clefting accrues. Recent data supporting a role for rare copy number variants (Lansdon et al., 2023) and how to incorporate other genomic variants, including those in non-coding regions (Zieger et al., 2023), will require additional data and validation through analytic trials. But as some individuals with a positive family history will have questions about risks consideration should be given to sequencing studies to identify variants that might suggest higher than what epidemiologic recurrence risks alone would support.

## Data Availability

Sequence and phenotype data is available from the Database of Genotypes and Phenotypes (dbGaP), study accession: phs001675.v1.p1.

## Acknowledgements

We would like to express our gratitude to the families involved in this study for their participation and we would like to thank our collaborators who have helped develop and produce this work. This study was supported with funding from the National Institutes of Health (X01-HG010012, T32-GM008490, R00-DE025060, R01-DE016148, R01-DE011931, R01-DE09886, R01-DE012472, P50-DE016215, R37-DE008559, R01-DE014667, R01-DD000295) and the Howard Hughes Medical Institute through the James H. Gilliam Fellowships for Advanced Study program. Sequencing services were provided by the Center for Inherited Disease Research (CIDR). CIDR is fully funded through a federal contract from the National Institutes of Health to The Johns Hopkins University, contract number HHSN268201700006I.

## SUPPLEMENTAL DATA

**Supplemental Table 1. Demographics of Study Cohort**.

**Supplemental Table 2. Variants of Interest in Multiplex OFC Families**.

